# Public health benefits of shifting from inpatient to outpatient TB care in Eastern Europe: optimising TB investments in Belarus, the Republic of Moldova, and Romania

**DOI:** 10.1101/2022.08.16.22278850

**Authors:** Sherrie L Kelly, Gerard Joseph Abou Jaoude, Tom Palmer, Jolene Skordis, Hassan Haghparast-Bidgoli, Lara Goscé, Sarah J Jarvis, David J Kedziora, Romesh Abeysuriya, Clemens Benedikt, Nicole Fraser-Hurt, Zara Shubber, Nejma Cheikh, Stela Bivol, Anna Roberts, David P Wilson, Rowan Martin-Hughes

**Affiliations:** Burnet Institute, Melbourne, Australia; Institute for Global Health, University College London, London, UK; Applied Sciences, Babylon Health, London, UK; Complex Adaptive Systems Lab, University of Technology Sydney, Sydney, Australia; World Bank Group, Washington, DC, USA; World Health Organization, Regional Office for Europe, Copenhagen, Denmark

**Author notes:** Corresponding author: Dr Rowan Martin-Hughes.

**Keywords:** tuberculosis, TB, outpatient care, ambulatory care, Eastern Europe

## Abstract

**Background:** High rates of drug-resistant tuberculosis (DR TB) continue to threaten public health, especially in Eastern Europe. Costs for treating DR TB are substantially higher than treating drug-susceptible TB, and higher yet if DR TB services are delivered in hospital. Therefore, countries are encouraged to transition from inpatient to ambulatory-focused TB care, which has been shown to have non-inferior health outcomes.

**Methods:** Allocative efficiency analyses were conducted for three countries in Eastern Europe, Belarus, the Republic of Moldova, and Romania to minimise a combination of active TB cases, prevalence of active TB, and TB-related deaths by 2035. These mathematical optimisations were carried out using Optima TB, a dynamical compartmental model of TB transmission. The focus of this study was to project the health and financial gains that could be realised if TB service delivery shifted from hospital to ambulatory-based care.

**Findings:** These analyses show that transitioning from inpatient to ambulatory TB care could reduce treatment costs by 5%−31% or almost 35 million US dollars across these three countries without affecting the quality of care. Improved TB outcomes could be achieved without additional spending by reinvesting these potential savings in cost-effective prevention and diagnosis interventions.

**Conclusions:** National governments should examine barriers delaying the adoption of outpatient DR TB care and consider the lost opportunities caused by delays in switching to more efficient and effective treatment modes.

## Introduction

In most Eastern European countries, the TB care model is based on legacy systems of inpatient care with injectable DR TB treatment. Historical models used long-term quarantine and allowed TB patients to recover over time, as these models were developed when effective DR TB drugs were not available and MDR TB did not exist (1). In Eastern Europe there has been a slow move towards outpatient TB care, particularly for countries with centralised economies. This delay may partly stem from legacy financing of TB sanatoriums and bed-based payment modalities. As a result, in 2019 17% (95% UI 16−18%) of new TB cases in Europe were multidrug-/rifampicin-resistant (MDR/RR) compared with only 3.3% (95% UI 2.4−4.4%) worldwide. Similarly, in Europe 52% (95% UI 45−59%) of cases were previously treated for MDR/RR TB versus only 18% (95% UI 9.7−27%) globally (2).

World Health Organization (WHO) guidelines issued in 2011 recommended investment in “systems that primarily employ ambulatory models of care to manage patients with drug-resistant TB over others based mainly on hospitalization” (3). Based on evidence from observational studies in Estonia, the Russian Federation, Peru, and the Philippines these guidelines were updated in 2019 (4) and 2020 (5) and maintain the recommendation to treat drug-resistant TB using primarily ambulatory models of care (i.e. services administered in a healthcare facility outside of hospital or in the community including home-based care provided by a community worker). The 2020 WHO guideline update on DR TB treatment states that “despite the limitations in the data available, there was no evidence that was in conflict with the recommendation, and which indicated that treatment in a hospital-focused model leads to a more favourable treatment outcome” (5). Moreover, a systematic review by Ho and colleagues that sourced evidence from a wide range of health settings provided additional support for ambulatory care over hospital-focused models of care for patients infected with multidrug-resistant TB (6).

In most Eastern European countries, the TB care model is based on legacy systems of inpatient care with injectable DR TB treatment. Historical models used long-term quarantine and allowed TB patients to recover over time, as they were developed at a time when effective DR TB drugs were not available and MDR TB did not exist (1). Particularly once effective DR TB drugs became available, the emergence and persistence of DR TB is a direct consequence of failings in the health care system (1). However, regardless of the availability of effective DR TB drug regimens and updated global health guidance, lengthy inpatient care models persist in most Eastern European countries and barriers to adopting outpatient DR TB treatment models still exist. These may involve health financing mechanisms that reimburse based on hospital bed occupancy rates for DR TB care or financing frameworks based on a restrictive line item budget making purchaser-provider split impossible. To overcome these types of barriers, solutions for health financing reform should consider results-based reimbursement and financing frameworks should allow for a more flexible global budget (1).

Avoiding hospital admissions, particularly to facilities with inadequate mechanisms for infection control, has been a key factor in reducing the risk of nosocomial transmission including the spread of TB and DR TB (1). Moreover, with the onset of the COVID-19 pandemic in early 2020, there has been an accelerated move to outpatient TB care to avoid the risk of SARS-CoV-2 infection for services sought in hospital. This shift is anticipated to continue in line with recommendations from the three country studies considered here. While some provision for inpatient TB care will likely remain to deliver specialised care for those with particularly complex cases, this shift to outpatient care is anticipated to continue.

While the overall burden of TB in Eastern Europe has declined in the last two decades, the incidence of drug-resistant TB has increased. In Belarus, the Republic of Moldova, and Romania, TB incidence, active TB prevalence, and TB-related deaths declined between 2000 and 2015, while the relative share of multidrug-resistant (MDR) and extensively drug-resistant (XDR) TB increased or continued over this period or at least did not decrease in these countries (key country information listed in Table 1). It is worth keeping in mind that the capacity to detect DR TB has significantly improved over the past decade (7). This has mainly been due to improved access to diagnostic technologies and rollout of rapid molecular diagnostics in high-burden countries.

**Table 1.** Key tuberculosis epidemic, finance, and programme information for Belarus, the Republic of Moldova, and Romania.

| Key category | Belarus<br>(8, 9) | Moldova (Republic of)<br>(10-12) | Romania<br>(13, 14) |
| --- | --- | --- | --- |
| Reporting year | 2015 | 2016 | 2018 |
| WHO classification | High MDR TB burden | High MDR TB burden | Not high TB burden |
| Est. MDR/RR TB incidence (1000s) | 3.5 (2.8–4.2) | 2.3 (1.9–2.6) | 0.71 (0.56–0.88) |
| TB financing |  |  |  |
| Total spending | US\$50.8 million | US\$17.7 million | US\$131.5 million |
| Spending for TB treatment | US\$47.1 million | US\$13.4 million | US\$100.6 million |
| *Domestic* |  |  |  |
| % of total | 89% | 77% | 49% |
| Description | Over 75% to hospital care | Not reported | Not reported |
| *International* |  |  |  |
| % of total | 10% | 23% | 11% |
| Description | Nearly 60% to ambulatory care | Not reported | Not reported |
| *Private* |  |  |  |
| % of total | <1% | None reported | 40% |
| Description | Primarily for ambulatory care | Not applicable | Not reported |
| National TB policy | WHO-recommended rapid diagnostic as initial test for all presumed to have TB (92% compliance) and universal access to drug susceptibility testing (100% compliance) (2) | National TB policy for high MDR TB burden, indicating WRD as the initial diagnostic test for people presumed to have TB (11) | Not available |

Entries were current at the time of each country analysis. MDR/RR= multidrug-/rifampicin-resistant. WRD=WHO-recommended rapid diagnostic.

In this study we focus on three case study countries in Eastern Europe where TB outpatient care programs have been defined. We compare optimised outcomes based on the savings gained from shifting to less expensive, safer, but equally effective outpatient TB care. For these countries we estimated how many more people could be reached each year with TB care (i.e. standard treatment) if savings from switching to outpatient care were cost-effectively reinvested in TB interventions.

These studies examine a reduction in unnecessary hospitalisation in-line with global DR TB care guidelines (4, 5), but do not remove hospitalisation entirely. There is plausibly no clinical benefit of DR TB treatment delivery in hospital for the majority of cases (unless hospitalisation is necessary where clinically indicated for the minority of cases) compared with outpatient primary care. The motivation to transition from inpatient to outpatient DR TB care is to not only save costs for the health system and for patients and their families (including lost income due to hospital stays estimated at 60% of out-of-pocket expenses as reported in a 2014 review in low- and middle-income countries (15)), but also to reduce the risk of nosocomial transmission.

From 2014 to 2018, 14 of the 15 countries in Eastern Europe and central Asia (EECA) reduced their number of bed days per TB patient. Overall Belarus was able to reduce their overall bed days for treatment by over 20% from 2015 to 2018. The number of hospital bed days per MDR or XDR patient per year were reduced from 120 to 115 days, although this is largely in line with the reduction in the number of TB cases that were projected. Romania was able to reduce their bed days per patient by 11% over this period with the relative size of the reduction influenced by both the percentage of TB patients hospitalised and the average length of stay if hospitalised (14).

## Materials and Methods

### Model and optimisation studies

Mathematical optimisation of TB spending was conducted using Optima TB, a dynamic population-based model of TB transmission fully described in (15) for three countries in Eastern Europe: Belarus, the Republic of Moldova, and Romania. All studies were conducted in collaboration with local stakeholders. An analysis was conducted in 2016−2017 for Belarus with a full description of the methodology provided in (8). Analyses were conducted in 2017−2018 for the Republic of Moldova as described in (10) and Romania as described in (13). The objective for these studies was to identify the most cost-effective resource allocation across existing and prospective TB diagnosis and treatment modalities to minimise a combination of active TB cases, prevalence of active TB, and TB-related deaths by 2035. A primary focus was to determine the health benefits and savings that could be gained by shifting from inpatient to outpatient TB care. This approach aligns with targets established in the National Tuberculosis Programme strategic plans for the countries considered.

### TB treatment modalities

Table 2 lists the outpatient-focused interventions considered for each country study. Duration of inpatient and outpatient TB treatment by modality for each country are shown in Supporting Information Tables S1−S3.

**Table 2.** Outpatient-focused treatment modalities considered in the modelling studies for Belarus, the Republic of Moldova, and Romania.

| **Outpatient-focused TB interventions** | **Belarus (8)** | **Moldova (Republic of) (10)** | **Romania (13)** |
| --- | --- | --- | --- |
| DS treatment | Standard and incentivised (incorporates financial incentives) modalities | Considered | Standard and directly observed therapy, short course (DOTS) |
| Short-course MDR treatment | Standard and incentivised modalities | Not considered | Standardised under direct (DOTS) and supportive observation |
| Long-course MDR treatment | Standard and incentivised modalities | MDR classic and MDR plus | Standard with and without Bedaquiline or delamanid |
| XDR treatment | Standard and incentivised ambulatory modalities | pre-XDR and XDR standard and with the addition of new drugs | Standard without Bedaquiline or delamanid |
| New and repurposed XDR drugs | Incentivised ambulatory modalities included the addition of Bedaquiline, clofazimine, and linezolid | New drugs included Bedaquiline, linezolid, imipenem/cilastatin, and amoxicillin/clavulanic acid | Standard with the addition of Bedaquiline or delamanid |

DS=drug-susceptible. MDR=multidrug-resistant. XDR=extensively drug-resistant.

### Study data and costing

For each country study, epidemiological, program, and cost data were collected by in-country experts and modellers in collaboration with international stakeholders. Literature reviews were also conducted to inform model parameters for each country, including intervention effectiveness and to support assumptions that had to be made as described in each country report (8, 10, 13). TB costing exercises were carried out for each country with costing data shown in Supporting Information Table S4 for Belarus, Table S2 for Moldova, and Table S5 for Romania. Costs represent the full cost of delivering a given intervention including commodities, delivery costs, staff time and TB-related costs outside the TB programme, such as TB-related hospitalisation by treatment modality and facility costs. For TB treatment interventions by modality (DS, MDR, and XDR), drug regiment costs (full course), inpatient costs, outpatient and directly observed therapy short course (DOTS) costs, and other related costs were included.

For Belarus, baseline spending by TB intervention and treatment type was established using the 2015 expenditures from WHO national health sub-accounts. Since which were triangulated with unit costs from other countries and international costing data to establish estimated spending by intervention as shown in the Supporting Information Table S4. TB drug cost per course of treatment by modality were including domestic and international donor funding (the Global Fund) for the 2015 calendar year. For Moldova, 2016 expenditure data sourced from WHO databases and reports, national TB reports for the WHO and the Ministry of Health, and National TB Programme records were triangulated with other unit cost data to establish estimated spending by intervention (Supporting Information Table S2). Costs were calculated considering the number registered TB patients, annualised costs, with other costs accounting for adverse drug reaction monitoring costs including costs of tests (such as audiometry, thyroid function, liver functioning, and electrocardiogram), which were mainly associated with drug resistant cases of TB. For Romania, costs for all treatment programs were estimated using a ‘bottom-up’ approach, based on average daily costs from hospital data. An average cost per ambulatory interaction was also derived and applied to screening programs and outpatient treatment following the initial hospitalisation period. (Supporting Information Table S5).

### Model calibration and cost-functions

Country models were calibrated primarily to TB case notifications and registered TB deaths. Cost-functions representing the relationship between spending and coverage, and coverage and outcome were generated. Calibrations and cost functions were validated together with in-country stakeholders.

### Optimisation approach

Using each country model, allocative efficiency projections were simulated for the total TB budget including for prevention, diagnostic, and treatment interventions. The potential for expanded diagnosis through active case finding was informed by country stakeholders when setting the model constraints for each program, and it is assumed that all those diagnosed will be eligible to receive TB treatment. Optimisation solutions for each country that best met the defined objectives were selected. From these reallocations, optimised TB treatment program spending for hospital-focused and ambulatory-based care, as well as for other treatment interventions (palliative care, prison-based treatment) were compared with the latest reported treatment spending. As part of the total TB budget optimisation, if less expensive but equally effective ambulatory TB treatment interventions (with costs provided in Supporting Information Tables S2, S4, and S5) are determined to be more impactful in achieving defined objectives by 2035 than hospital-based treatment, then the model algorithm will relocate resources accordingly.

For this modelling analysis it was assumed that any savings from prioritising more cost-effective ambulatory TB services would be reinvested in TB programs, versus disbursed, at least in part, to other health areas. However, as reported at the time of the original analyses and explored through follow-up interviews with study country teams (conducted in 2021), there have been structural limitations with healthcare financing that have restricted opportunities to reinvest savings from one area of TB programming into another. These limitations should be examined. Nevertheless, whether governments decide to reinvest savings directly in TB programmes, in other areas of health, or in non-health related sectors, there are opportunities for the country to benefit. Therefore, any potential gains should be pursued and lost opportunities avoided.

### Outcomes

As part of optimising resource allocations for Belarus, Moldova, and Romania more cost-effective ambulatory treatment modalities should be prioritised. This will lead to cost savings, with the recommendation to reinvesting these savings to increase ambulatory treatment coverage. This also includes the earlier diagnosis of additional TB cases, which in turn will allow more people to receive treatment. The number of cumulative active TB cases and TB-related deaths that could be averted by 2035 were estimated. The reduction in the prevalence of active TB that could be achieved over this period was also projected. This analysis draws together common results and conclusions from TB budget impact studies for Belarus, Moldova, and Romania, with a focus on projected health and financial gains that could be realised by prioritise ambulatory TB care.

## Results

Moving from hospital-focused to ambulatory TB care would yield positive public health benefits in all three country settings. For Belarus, transitioning from the 2015 model of hospital focused TB care to ambulatory care could reduce TB treatment costs by nearly US$15 million or 31% by 2035 (Fig. 1). At the time of this analysis, it was projected that TB cases in Belarus would decline in the future, which would result in fewer people needing TB treatment. Immediate savings from transitioning from involuntary isolation and other hospital-focused treatment to outpatient care, as well as savings if new cases decline as projected meaning reduced need for treatment, should be reallocated to higher impact program interventions and delivery solutions. These include providing incentives to improve patient adherence and ambulatory care outreach, procuring new, more efficacious drug regimens for MDR and XDR TB, scaling up rapid molecular diagnostics, enhancing active case finding among high-risk populations, and enhancing contact tracing.

**Fig. 1.**
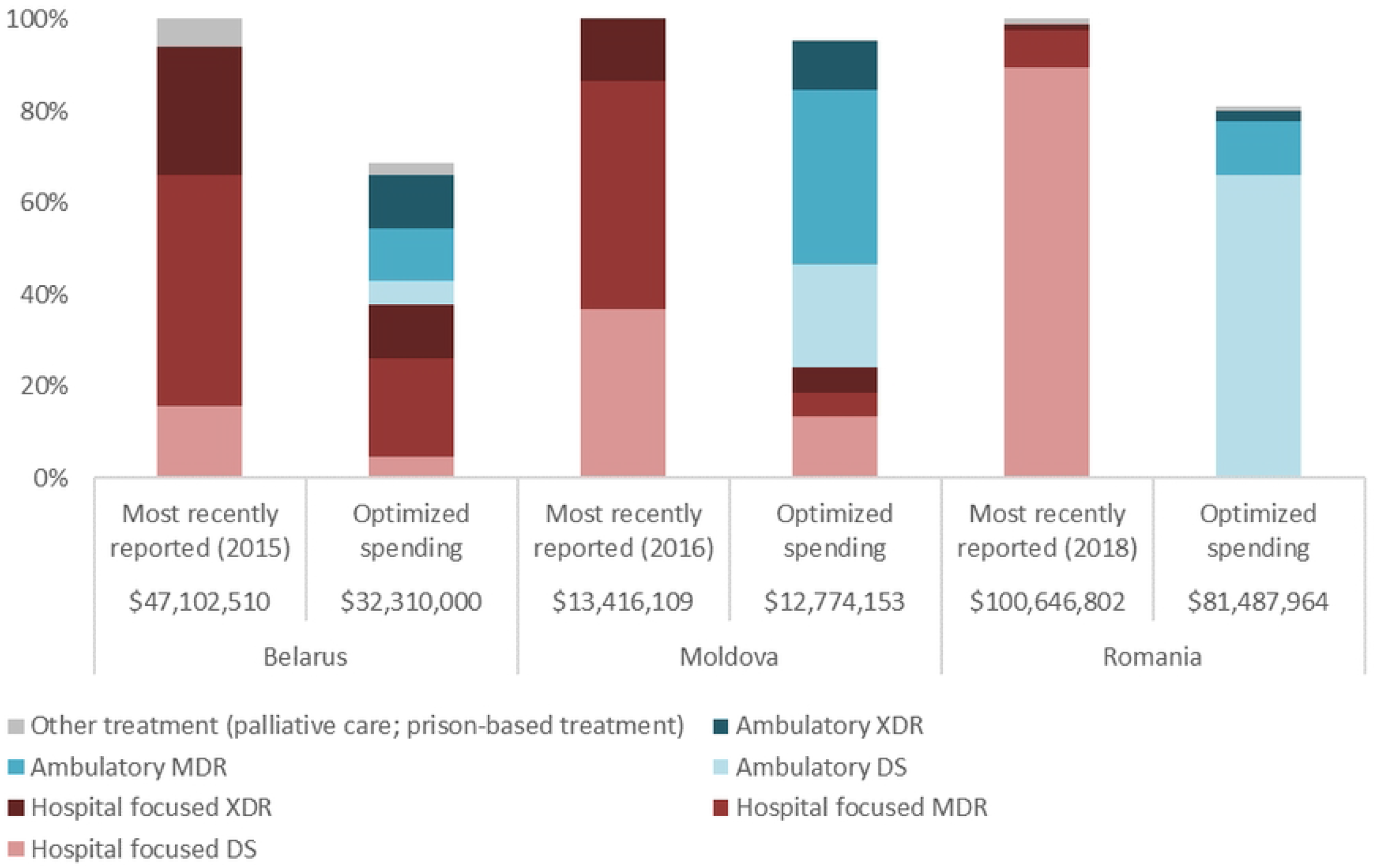
Optimised annual tuberculosis TB treatment allocations relative to the most recently reported spending by treatment modality represented as a percentage of total TB programme spending (for a given reporting year) for Belarus (8), Moldova (10), and Romania (13). TB treatment interventions include hospital-focused and ambulatory-based care for DS, MDR, and XDR TB, as well as other treatment interventions (palliative care, prison-based treatment). Values for the most recently reported annual TB treatment budget and optimised resource allocations for all TB treatment interventions are indicated below their respective bars for each country. Spending values provided in Euros for the Moldova (10) and Romania (13) modelling studies were converted to USD corresponding to the year spent (at the time of original analyses). Spending was provided in USD for Belarus (8). DS=drug-susceptible. MDR=multidrug-resistant. XDR=extensively drug-resistant.

For example, in Belarus, there were 264 patients treated in hospital for DR TB in 2015. These modalities had the highest unit costs, $21,482 for a full long course of MDR treatment and $28,840 for XDR treatment in 2015 USD. US$16.6 million was spent on these modalities accounting for 26.8% of all TB-related spending in that year. As well, this transition to ambulatory care resulted in reductions in duration of hospital stay from 60 to 14 days for drug susceptible (DS) TB treatment, 210 for MDR to 45 days for long-regimen and 30 for short-regimen, and 270 to 60 for XDR TB care.

Within the same total national TB budgets for each country, annual TB treatment coverage values under total TB resource optimisation to best achieve objective targets through to 2035, as well as the most recently reported coverage values are shown below the respective bars in Fig. 2. Coverage values by type of TB treatment (DS, MDR, and XDR) are also represented graphically as a percentage of treatment need. For Moldova and Romania, treatment coverage under optimised allocation would surpass the need most recently reported (at the time of analysis), 121% and 104%, respectively. Uniquely for Belarus, since TB cases are projected to decline in the future, meaning less people would need treatment, over 30% fewer people are estimated to need coverage each year for DS treatment under optimised allocation. Coverage for drug-resistant TB modalities are predicted to marginally increase with cost-effective reallocation for this country.

**Fig. 2.**
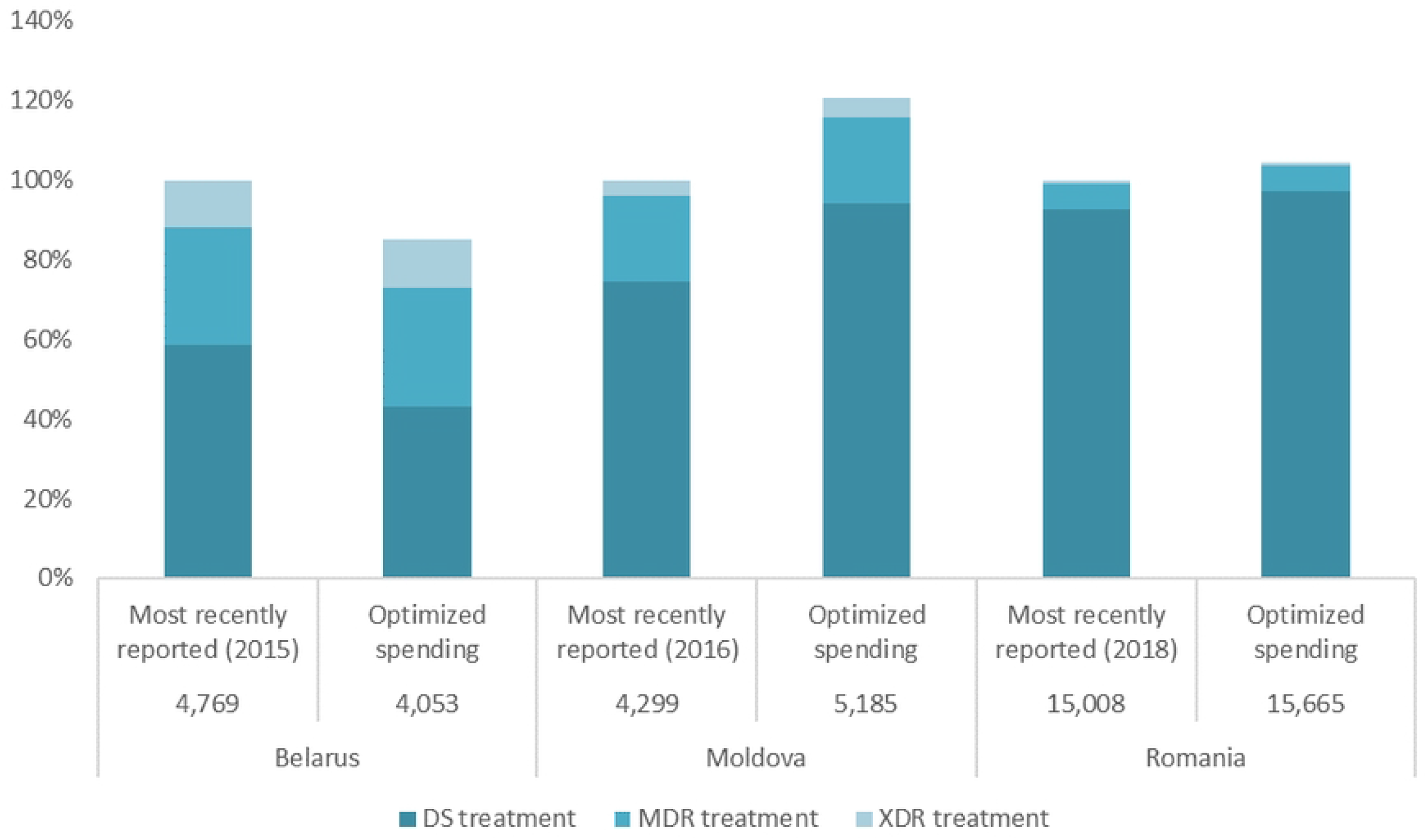
Annual TB treatment coverage under optimised allocation of resources for all TB interventions compared with the most recently reported treatment coverage for Belarus (8), Moldova (10), and Romania (13). The annual number of people on TB treatment most recently reported and under optimised allocation are indicated below the respective bars for each country. DS=drug-susceptible. MDR=multidrug-resistant. XDR=extensively drug-resistant.

As part of the modelling analysis conducted for Moldova, it was estimated that prioritising ambulatory care could reduce treatment costs by an estimated 5%, potentially freeing up approximately US$0.6 million for reallocation to higher impact interventions including reinvestment to increase treatment coverage. The largest relative proportion of this saving comes from MDR and XDR TB treatment programs that have the longest duration of treatment programs at a duration of 18 to 24 months. Lengthy hospitalisation is the primary cost driver of the TB response in Moldova. Based on national program records, the duration of hospitalisation could be reduced substantially from 40 to 14 days on average for DS TB treatment to align with international practice. Hospitalisation for drug-resistant TB treatment could be reduced from 45 days for long-regimen MDR TB and to 30 days for short-regimen, and from between 127 and 195 days to 60 days for XDR (10). Reduced hospitalisation for XDR TB cases (excluding pre-XDR) would allow for increasing coverage by up to 153%, which would in principle allow nearly every person with XDR TB who is aware of their status to be on treatment with new Bedaquiline-based pre-XDR and XDR regimens where eligible or standard regimens where not available. It was recommended that any resources freed up by changing treatment modalities should be invested in selected higher impact interventions and delivery solutions. These include provision of incentives for providers of ambulatory TB care, procurement of new, more efficacious drug regimens for MDR TB and XDR TB, scale up of rapid molecular diagnostics, enhanced active case finding among high-risk populations, and enhanced contact tracing (10).

Finally, the analysis for Romania also confirmed that transitioning to ambulatory treatment after a reduced initial hospitalisation could reduce the cost of TB treatment by US$19.2 million, a 19% reduction in current expenditure. Reductions in duration were as follows, from 67 to 21 days for DS TB, from 180 days to 30−60 days for MDR TB, and from 270 days to 120−180 days for XDR TB, including the use of direct observed therapy, short course (DOTS) where appropriate (13).

If resources for TB were optimally reallocated from 2015 to 2035 for Belarus, Moldova, and Romania, including prioritising less expensive but equally effective ambulatory TB care (therefore more cost-effective) over hospital-based care, and assuming these savings remained in the TB budget and were optimally reinvested across TB interventions, then new active TB infections could be reduced by 9% in Moldova (1% in Romania and 7% in Belarus), active TB prevalence per 100,000 reduced by 44% in Moldova (5% in Belarus and 27% in Romania), and TB-related deaths reduced by 48% in Moldova (5% in Belarus and 21% in Romania) over this period (Fig. 3 with corresponding estimates reported in Supporting Information Tables S6−S8). Focusing on maximising TB outcomes, not considering potential benefits for other areas of health, modelling shows these savings should be optimally reinvested in TB prevention, diagnosis, and ambulatory treatment interventions to increase treatment coverage. In each country, increased investment in active case finding (particularly in high incidence areas and to target high-risk groups) and prevention was projected to lead to rapid decreases in the prevalence of active TB prevalence and TB-related mortality, but the high burden of latent TB means that new active TB infections are projected to decline more slowly.

**Fig. 3.**
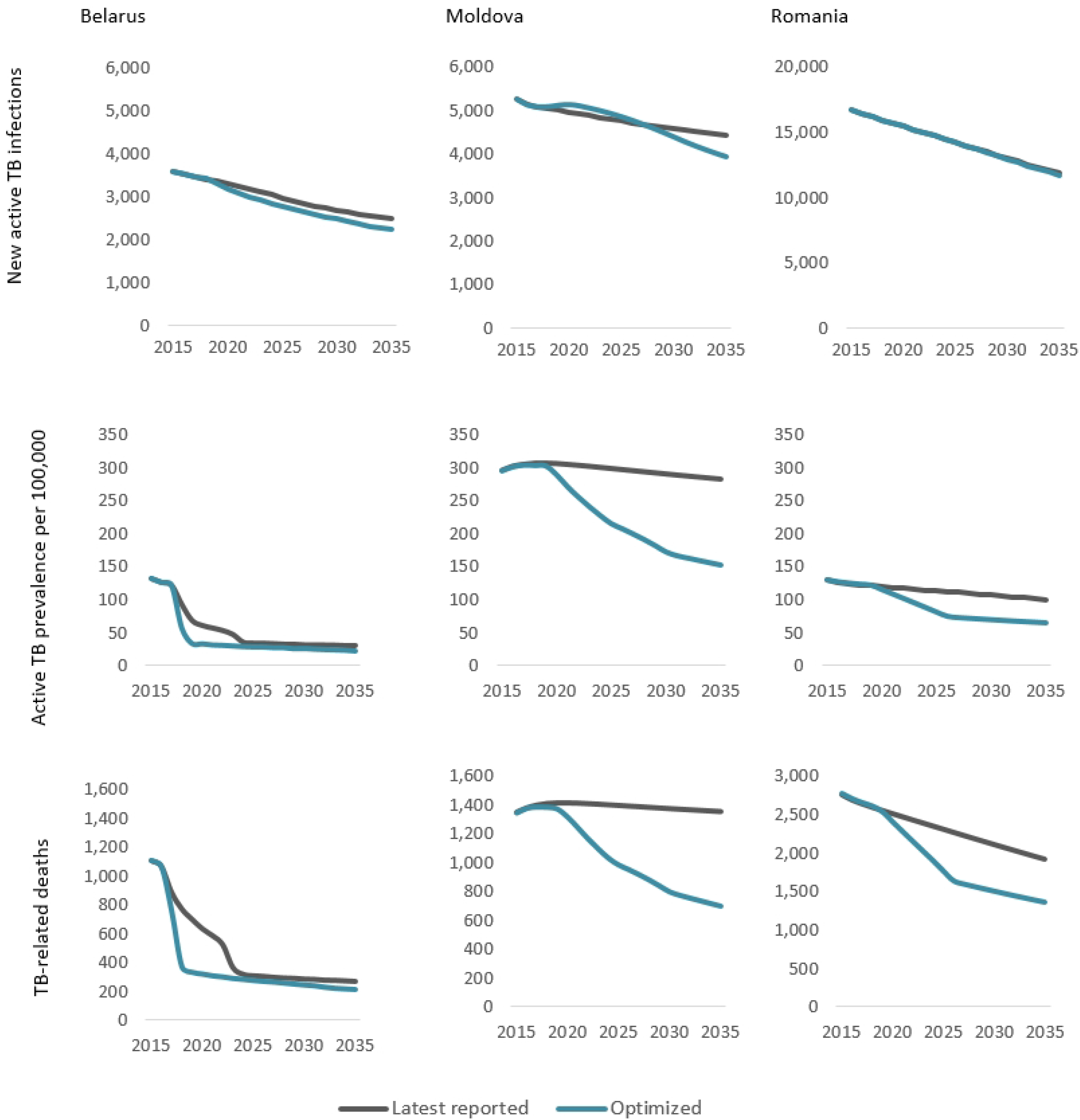
Projected reductions in new active TB infections, active TB prevalence, and TB-related deaths under optimal allocation of treatment resources from 2015 to 2035 for Belarus (8), Moldova (10), and Romania (13). This includes prioritisation of less expensive ambulatory care and resulting savings being optimally reinvested in TB prevention, diagnosis, and additional ambulatory treatment.

## Discussion

Importantly, evidence suggests that ambulatory care for those with drug-resistant TB infection has at least the same treatment outcome as hospital-focused care (5). Moreover, Williams and colleagues observed better MDR TB treatment success for outpatient treatment compared with traditional hospitalisation for nine countries in Africa, Asia, and Eastern Europe (16), as was also reported for the Republic of Macedonia (17). Similarly, Ho and colleagues reported that success was more likely for outpatient care from eight studies in Africa, Asia, and the USA (6). The 2019 WHO guidelines on DR TB conditionally recommend that “patients with MDR TB should be treated using mainly ambulatory care rather than models of care based principally on hospitalization” (4). Despite low quality evidence from observational studies used to inform the 2020 updated WHO guidelines on DR TB care, the guidelines state that “there was no evidence that was in conflict with the recommendation, and which indicated that treatment in a hospital-focused model of care leads to a more favourable treatment outcome” (5). Here we demonstrate that transitioning from hospital-to ambulatory-based DR TB treatment could yield savings of 31%, 5%, and 19% in Belarus, Moldova, and Romania, respectively, while achieving at least comparable projected treatment outcomes (Figs. 2 and 3). It is recommended that these savings be optimally reinvested in TB prevention, diagnosis, and outpatient treatment to achieve increased treatment coverage and further health gains.

As part of reinvesting savings to increase treatment coverage, options for increasing treatment adherence such as abbreviated treatment regimens, expanded patient incentives, and community support interventions should be explored and benefits tracked to inform future analyses. This could not be assessed in these studies due to paucity of data at the time of analysis.

An important benefit of conducting these country studies came from the extensive costing exercises that were undertaken. Collecting costing components and deriving cost per treatment course by TB treatment modality, DS, MDR, and XDR, and as well as whether delivered in-hospital or at outpatient care facilities or in the community then allowed comparison of potential saving and health gains that could be realised by prioritising ambulatory care. However, prioritising TB treatment delivery from inpatient to outpatient care (18-20) will involve more than decision-making on funding reallocation. This transition will require shifting emphases in care models through changes in clinical guidelines, changes in how funding flows to facilities, or through incentives. This may also include task shifting and other changes to human resourcing, as well as changes in demand-side expectations for hospital versus ambulatory care. Lastly, in many settings TB care financing reform may not be a short-term process and may require different approaches and timeframes.

The modelling study in Belarus provided evidence that led to a recommendation to strengthen ambulatory care through incentives to improve healthworker outreach support and patient adherence. It was suggested that this recommendation be fulfilled using a combination of delivery solutions, which are likely to improve treatment outcomes. It is acknowledged that enhanced ambulatory care requires a reform of tuberculosis care financing to replace bed-based payment with outcomes-based financing. In Moldova, as reported in the 2020 WHO Global TB report, “It is also evident that some EECA countries have markedly reduced their use of hospitalisation and have changed their model of care for people with drug-susceptible TB”. As noted previously, from 2014 to 2018, 14 of the 15 EECA countries reduced the number of bed days per person (14). The size of the reduction, which is influenced by the percentage of people with drug-susceptible TB who are hospitalised and the average length of stay if hospitalised, ranged from 21% in the Republic of Moldova to 81% in the Russian Federation. As such, new active case-finding modalities were being introduced as of 2019. Mobile outreach vans were being piloted to target high-risk populations, with the aim of ensuring early diagnosis and treatment for people who are typically hard to reach. A separate study is underway together with national stakeholders to assess whether recommendations from these modeling studies have been adopted, how they have been implemented, and what benefits may have been gained as a result, as well as lessons learnt. This new study will include these countries in Eastern Europe, but other country studies and disease areas will also be included.

The COVID-19 pandemic has resulted in a shift to outpatient care to avoid the risk of SARS-CoV-2 infection. This was achieved through technical advances including telehealth, video supported treatment, and other lower contact service delivery approaches. Many of these innovations were in place before the pandemic, but the pandemic prompted the transition to utilise these modalities making it more convenient and decreasing the burden for both patients and providers in ambulatory settings. It is anticipated that many of these care options will continue, even once the need for the COVID response lessens. Given the potential gains from furthering shift towards outpatient care, as estimated here, it would be advantageous for TB programme planners to continue incorporating this shift in service delivery into ongoing TB response plans.

Following global guidelines to transition away from hospital-based to outpatient DR TB care (5) there are other benefits beyond cost savings, which were not captured in this analysis. Other benefits include reduced nosocomial transmission-related health systems costs, cost (direct and indirect) to the patient, as well as reduction in infection risk, and stigma surrounding access to longer-term hospital care. It may also be worth exploring the cost-effectiveness of integrating DR TB care services with other health programs, particularly those delivered more readily in ambulatory care settings, such as mental health services and alcohol cessation support. One such example is for people coinfected with TB and HIV; co-treatment could be decentralised through ambulatory care and therefore be more patient-centered, could result in healthcare cost savings, reduced loss in income through avoided hospital stays, and other benefits (21).

An international systematic review of the evidence supports the assumption that ambulatory care could achieve current coverage levels in target populations (22). A meta-analysis of 540 articles reported no statistical difference for treatment outcome rates (success, death, default, and failure), between ambulatory and hospital-focused delivery of TB care. The review found that standard ambulatory care can be as effective as hospital-focused care (22). There is also evidence to suggest that ambulatory care that is enhanced by specific incentives might be more effective than standard ambulatory care. A Cochrane review suggested that ambulatory care coupled with cash incentives for patients may be more effective than non-incentivised ambulatory care, particularly among high-risk groups (23). A WHO review of evidence also suggests improvements in treatment adherence through food and financial support as well as TB care enhanced through a mix of interventions (24). Considerations around a complete shift from hospital-focused to ambulatory care are that comorbidities, including alcohol use disorder, and coinfection with HIV (non-homogeneous), are also common in this region. In future, more complex cases will likely still need at least some hospitalised care.

As part of health reforms that emphasise people-centred care and favour results-based financing (1), countries are encouraged to adopt care models that replace inpatient care for injectable DR TB treatment with ambulatory care with oral regimens for drug-susceptible and drug-resistant TB that have fewer side effects and favour decentralised TB care models (4, 5). Although it was not the focus of this study, in other settings ambulatory care has also been shown to drastically reduce out-of-pocket expense for people receiving TB treatment (25). As a follow-on to these studies, most countries in Eastern Europe are currently transitioning towards ambulatory TB care (26).

## Data Availability

Country-specific model data are available through respective country-specific reports as cited in this study.

## Acknowledgements

The authors gratefully acknowledge contributors to the country studies that formed the basis of this work, including the following representatives: Dzmitry Klimuk, Alena Skrahina, Henadz Hurevich (Republican Scientific and Practice Centre for Pulmonology and Tuberculosis, Belarus); Inna Nekrasova, Marina Sachek, Vassily Akulov (Republican Scientific and Practice Centre for Medical Technologies, Belarus); Alena Tkatcheva (Ministry of Health, Belarus); Dragutan Cristina, Lilia Gantea (Ministry of Health, Labour, Social Protection, Moldova); Rita Seicas (Center for Health Policies and Studies, Moldova); Victoria Petrica (Project Coordination and Implementation Unit, Moldova); Valeriu Sava (SDC, Moldova); Sofia Alexandru, Diana Condratchi, Andrei Corlateanu, Valeriu Crudu, Nicolae Nalivaico, Valentina Vilc, Liudmila Marandici (‘Chiril Draganiuc’ Institute of Phthisiopneumology/National Tuberculosis Programme, Moldova); Munteanu Ioana (National Tuberculosis Programme, Romania); Moldova Adriana Socaci, Beatrice Mahler-Boca, Domnica Ioana Chiotan, Gilda Popescu, Mihaela Stefan Nicoleta Cioran (Romania National Institute of Pulmonology); Amalia Serban, Ana-Maria Ciobanu, Costin Iliuta, Mihaela Bardos (Ministry of Health, Romania); Dana Farcasanu, Daniel Ciurea (Center for Health Policies and Services, Romania); Fidelie Kalambayi (Romanian Angel Appeal); Mihnea Dosius (Romania National School of Public Health); Viatcheslav Grankov Valentin Rusovich, Andrew Siroka, Cassandra Butu (World Health Organization); David Kokiashvili George Sakvarelidze, Sandra Irbe (Global Fund); Azfar Hussain, Janka Petravic, Cliff Kerr (Burnet Institute); Ibrahim Abubakar, Marius Nasta (University College London); and Feng Zhao, Marelize Görgens, Irina Oleinik, Irina Guban, Hanna Shvanok, Cristina Petcu, Huihui Wang, Jaime Nicolas Bayona Garcia, David Wilson (World Bank).

## Supporting information

**S1 File** Provides additional details on the TB modelling analyses conducted for Belarus, the Republic of Moldova, and Romania including duration and costs associated with different TB treatment modalities.

